# Use of Large Language Models by U.S. Adults to Support Exercise: A Survey Study

**DOI:** 10.64898/2026.05.01.26352211

**Authors:** Megan A. McVay, Selina Willfort, Danielle E. Jake-Schoffman, Bonnie Dorr, Amy J. Sheer, Kelsey Henry

## Abstract

**Background:** Large Language Model (LLM) chatbots are increasingly used for exercise and fitness topics, yet users’ experience with these tools remains understudied.

**Methods:** This study is a national survey of U.S. adults who have used an LLM chatbot for exercise-related topics in the past month. Participants answered questions about the exercise-related topics for which they used LLM chatbots, their perceptions of these chatbots’ value for exercise-related questions, and how chatbot use had changed their exercise behaviors and use of other exercise-related resources.

**Results:** Participants (n=258) were majority male (n=138, 53.5%) and white (n=146, 56.6%) with a mean age of 41.7 (SD=14.9) years. The most endorsed topics for LLM chatbot use were making an exercise plan (n=137, 53.1%), nutrition related to exercise (n=132, 51.2%), advice on amount of exercise (n=122, 47.3%), specific exercises to try (n=120, 46.5%), and motivation or emotional support for exercise (n=112, 43.4%). On average, participants endorsed high trust (M=4.0, SD=0.7; on 1-5 scale) and a moderate emotional bond (M=3.0, SD=1.3) with LLM chatbots. Most participants (n=140, 54.3%) reported that they increased their exercise due to LLM chatbot use (M=55.6 minutes increase). Some participants reported increases in use of other resources; e.g., gyms (26.4%), wearable technology (23.3%), and exercise questions to their healthcare providers (25.6%). Those who increased exercise with LLM chatbot use reported significantly higher trust (M=4.1 vs M=3.9) and emotional bond (M=3.2 vs M=2.6) with chatbots and more use for motivation/emotional support (70.5% vs 29.5%) compared to those who did not. Many participants also used LLM chatbots for nutrition and weight-related questions.

**Discussion:** LLM chatbots may meaningfully impact exercise-related behavior and resource use, warranting more rigorous causal research.

## Introduction

Large Language Model (LLM) chatbots are conversational agents that use generative artificial intelligence to respond to text prompts. LLM chatbots are easily accessible and are rapidly achieving a high level of population penetration among the US population; as of late 2024, three-quarters of US adults report awareness of LLM chatbots and nearly 40% report using LLM chatbots.^1^ ChatGPT, the most widely used LLM chatbot, alone was used by one-third of US adults as of early 2025.^2,3^ Use of LLM chatbots is expected to continue growing.^4^ Not surprisingly, LLM chatbots are also increasingly used by individuals as a source of information and guidance for their health. As of December 2025, 32% of US adults report using LLM chatbots for health information at least monthly.^5^ In an analysis of a million prompts inputted in the LLM chatbot Claude in early 2025, 1.5% of all queries were related to exercise/fitness and nutrition and a 2026 analysis of the LLM chatbot Co-Pilot found that 9% of all health questions were fitness-related.^6^

Physical inactivity is a significant contributor to chronic disease and mortality among US adults,^7,8^ and an important question is whether using LLM chatbots to receive exercise-related information, recommendations, or support can help individuals make positive changes in their physical activity. LLM chatbots have several features that make them likely to be successful in promoting behavior change such as increased exercise: they can provide tailored responses at the moment users seek support, are often perceived as non-judgmental,^9,10^ and may foster a strong working alliance with users.^11^ They may also address or inform on key behavior change mechanisms, such as stimulus control reinforcement management and social support.^12^ Studies have also shown that LLM chatbots produce health-related output that is rated more readable than human sources.^13^ Combined, these characteristics may result in potential for high engagement and successful behavior change.

Despite this promise of LLMs for health behavior change such as exercise, there are also reasons to believe that general-purpose LLMs will fail to be effective. Absence of human accountability and structured guidance may diminish the impact of LLM chatbots.^14^ Additionally, the ability to influence psychosocial behavior change mediators could be limited in general-purpose LLMs, especially if user inquiries are not focused on using the LLM to address psychosocial factors influencing exercise, such as motivation. An additional factor that may be important in the effectiveness of LLMs for promoting exercise is the users’ trust in the tool. A significant concern about LLMs is the presence of inaccuracies or false information (“hallucinations”), which could result in use of ineffective or unsafe strategies or lower trust in LLMs.^15,16^ Finally, individuals vary in the extent to which they feel a personal connection or emotional bond with the LLM chatbot they use.^17^ The extent to which users feel a bond may influence how impactful LLM chatbots are on exercise behaviors, similar to how bonds with therapist or coaches are linked to behavior change.^18–20^ To date, it is unknown if LLM chatbots are capable of impacting exercise or what factors might influence the effects of LLM chatbots on physical activity levels.

Another unanswered question about LLM chatbot use for exercise is how it will affect use of other tools and resources that are used for learning about and supporting physical activity. Potentially, LLM chatbot use could promote uptake of other resources by suggesting use of additional tools, from wearable devices to following of social media influencers. Yet LLM chatbots use may also displace use of some resources for fitness, for example, by reducing perceived need for personal trainers or preempting asking a healthcare provided questions about exercise.

To expand the currently limited understanding of how individuals use LLM chatbots for exercise and the effects of that use, we conducted a national survey among adults who report recent use of LLM chatbots for physical activity. This study makes three contributions: it characterizes how users are engaging LLM chatbots for exercise-related questions; it examines perceived trust, usefulness, and emotional bond in this context; and it provides early evidence on perceived changes in exercise behavior and related resource use. We addressed four questions: 1) What types of exercise-related topics are people seeking out LLM chatbots for?; 2) What are users’ perceptions of LLM chatbots usefulness and trustworthiness for exercise topics, and what is their extent of emotional bond with an LLM chatbot?; 3) How has using an LLM chatbot for physical activity topics changed use of other resources for exercise?; and 4) Do users perceive that the LLM chatbots have led them to change their exercise behaviors?

## Methods

### Study design and participants

This was a cross-sectional survey of adults reporting recent use of LLM chatbots for exercise-related topics. Eligibility criteria were age 18 or older, U.S. citizenship, ability to read English, and self-report of having used an LLM chatbot at least once in the past month and at least twice in the past year for an exercise- or fitness-related topic for the participant’s own use. Recruitment was conducted by the survey company YouGov among their proprietary participant panel. YouGov’s panel is comprised of over 1 million U.S. residents who are recruited through diverse methods including digital and print ads, partner sponsored solicitation, and social media. Participants are compensated by YouGov in points redeemable for gift cards. All data were collected in February 2026. This study was approved by the University of Florida Institutional Review Board. Participants were provided informed consent with waiver of documentation. LLM chatbots were used in preparation of this manuscript to assist with coding for data analyses.

### Study procedures and data quality measures

The specific purpose of the study was obscured during recruitment and screening in order to avoid deliberate misreporting. Survey panel members were invited to complete a survey via a generic invitation. Participants were asked to select how they use LLM chatbots from a list of 6 possible uses; if participants selected health/wellness, they were then asked to select from a list of 10 health-related topics with one being exercise/fitness. During screening, participants were also asked which LLM chatbots they have used, and the list included fake LLM names, which, if selected, excluded participants. The survey company has extensive quality control measures for their participant panels, including blocking of blacklisted emails, overactive IP addresses, and devices unable to pass a reCAPTCHA; regular evaluation of panel through open-ended questions and attention checks; and use of geo-location data. Additionally, respondents are removed from the sample by the survey company based on too rapid response time, providing irrelevant responses to open-ended questions, and evidence of duplicate responses based on IP address and cookies.

### Measures

Several items were created for this study by the study team to capture users’ experience with LLM chatbots. Prior to administering the present survey, these items were evaluated through Zoom-based cognitive interviews examining comprehensibility, clarity, and comprehensiveness with 6 participants from the target population; no additional interviews were conducted due to feedback saturation (i.e., no new concerns being raised). These interviews led to wording revisions and the addition of items related to exercise-topic use and use of other exercise-related resources.

The full survey is available in Supplemental Material. Items were included to characterize use of LLM chatbots, including questions about specific LLM chatbots used, frequency of use for general purposes and for exercise, if participants have paid for use, and time since first use for exercise. Participants were asked to indicate if they had used LLMs for a variety of exercise topics; this list was initially developed based on research team expertise and further refined during pilot testing with the target population. Survey respondents were also asked to share if use of LLM chatbots for exercise led to more, less, or no change in use of other exercise resources, such as personal trainers, YouTube videos, questions to their healthcare providers, and more. Participants were asked to indicate if they are exercising more minutes per week since using LLM chatbots, and if so, how many more minutes per week. They were further asked to report if their exercise had changed in any other way, and, if affirmative, an open-ended question asked them to state how it had changed. Perceptions of the information provided by the LLM chatbot were adapted from a scale developed and used previously by Camilleri & Camilleri.^21^ This measure included subscales related to perceived relevance, accuracy, trustworthiness, and usefulness. Items were adapted to ask specifically about information provided related to exercise (e.g., “I trust the content that is given by my AI Chatbot about exercise”), with response options ranging from 1 (strongly disagree) to 5 (strongly agree).

Emotional bond with LLM chatbot was assessed using the single item “I feel an emotional bond to my AI chatbot” with the same rating scale. Participants additionally completed the Standford Leisure Time Activity Categorical Item (LCAT), a validated measure of physical activity.^22^ Finally, participants also shared if they had communicated with an LLM chatbot about a variety of nutrition and weight-related topics, with this list of topics also developed by the study team and refined in pilot testing.

### Analyses

Descriptive analyses using means and standard deviation for continuous variables and frequencies for categorical variables were calculated. To compare those who reported an increase in exercise with those who did not, we conducted χ^2^ goodness of fit tests for categorical variables and independent samples t-tests for continuous variables, interpreting a p-value of <0.05 as statistically significant. Open-ended responses on types of activity changes were coded independently by two coders, who compared and reconciled codes after coding approximately 15% of responses, then again after 30%, 50%, and 100%. Quantitative data analyses were conducted in R studio version 2025 5.1. The deidentified data set and analytic code is available at the OSF project website (https://osf.io/rd9hq/overview?view_only=ad35b7b6c111498db0da83276c1e1fef).

## Results

### Participant characteristics

A total of n=5,320 participants had available screening data. Among these, n=2,491 (46.8%) reported any LLM use in the past month. Use of LLM chatbots for exercise at any point in prior 3 months was reported by n=283, equivalent to 5.3% of all screening respondents and 11.4% of all respondents who reported LLM chatbot use. A total of n=258 (4.8% of screening respondents) met full eligibility criteria, including use of LLM chatbots for exercise twice in the past year for themselves. Using our full screening sample, we compared those who met eligibility criteria with those who did not. Eligibility was more common among men (vs. women), those with a bachelor’s or higher degree (vs. those without), younger respondents, those with a higher income, and non-white respondents (see Supplemental Materials for data).

Characteristics of participants who were eligible for and completed the full survey are shown in Table 1. The sample was 53.5% male and 56.6% identified as white. The mean age was 41.7 (SD=14.9) years. A variety of education levels were represented, with 60.1% reporting a bachelor’s degree or higher educational attainment. A variety of levels of overall LLM chatbot use were reported, with 45% reporting at least daily use of LLM chatbots (See Table 2). Approximately 80% of users reported believing that they were at least moderately skilled in use of LLM chatbots.

**Table 1.** Participant characteristics (n=258).

|  | **N/M** | **%/SD** | **M** | **SD** |
| --- | --- | --- | --- | --- |
| Age | 41.7 | 14.9 | 41.7 | 14.9 |
| Gender |  |  |  |  |
| Man | 138 | 53.5 |  |  |
| Woman | 120 | 46.5 |  |  |
| Race |  |  |  |  |
| White | 146 | 56.6 |  |  |
| Black | 36 | 14.0 |  |  |
| Hispanic | 50 | 19.4 |  |  |
| Asian | 14 | 5.4 |  |  |
| Native American | 2 | 0.8 |  |  |
| Multi-racial | 7 | 2.7 |  |  |
| Other | 3 | 1.2 |  |  |
| Hispanic | 63 | 24.4 |  |  |
| Highest education |  |  |  |  |
| No high school | 4 | 1.6 |  |  |
| High school graduate | 57 | 22.1 |  |  |
| Some college | 40 | 15.5 |  |  |
| 2 year degree | 25 | 9.7 |  |  |
| 4 year degree | 77 | 29.8 |  |  |
| Post-graduate | 55 | 21.3 |  |  |
| Financial security |  |  |  |  |
| Very low | 20 | 7.8 |  |  |
| Low | 49 | 19.0 |  |  |
| Medium | 89 | 34.5 |  |  |
| High | 100 | 38.8 |  |  |
| Employment |  |  |  |  |
| Work full-time | 141 | 54.7 |  |  |
| Work part-time | 35 | 13.6 |  |  |
| Unemployed, disabled, student | 49 | 19.0 |  |  |
| Retired | 33 | 12.8 |  |  |
| Physical activity level<sup>a</sup> |  |  |  |  |
| Inactive | 9 | 3.5 |  |  |
| Light 1-2x/week | 54 | 20.9 |  |  |
| Moderate 3x/week | 76 | 29.5 |  |  |
| Moderate >=5x/week | 64 | 24.8 |  |  |
| Vigorous 3x week | 32 | 12.4 |  |  |
| Vigorous >= 5x/week | 23 | 8.9 |  |  |
| BMI, m/kg<sup>2</sup> |  |  | 26.1 | 6.5 |
| BMI category |  |  |  |  |
| <18 kg/m<sup>2</sup> | 5 | 1.9 |  |  |
| 18-24.9 kg/m<sup>2</sup> | 127 | 49.2 |  |  |
| 25-29.9 kg/m<sup>2</sup> | 77 | 29.8 |  |  |
| >30 kg/m<sup>2</sup> | 49 | 18.9 |  |  |
| Tried losing weight in past 3 months | 151 | 58.5 |  |  |

a. From Stanford Leisure-Time Activity Categorical Item (L-CAT).

**Table 2.** LLM chatbot use characteristics of study participants (n=258).

|  | N | % | M | SD |
| --- | --- | --- | --- | --- |
| LLM chatbot general use frequency (past 3 months) |  |  |  |  |
| Less than weekly | 20 | 7.8 |  |  |
| Weekly | 35 | 13.6 |  |  |
| Several times a week | 86 | 33.3 |  |  |
| Daily | 21 | 8.1 |  |  |
| Several times a day | 66 | 25.6 |  |  |
| Almost constantly | 30 | 11.6 |  |  |
| Perception of skill in use of LLM chatbot |  |  |  |  |
| Very/extremely skilled | 106 | 41.1 |  |  |
| Moderately skilled | 100 | 38.8 |  |  |
| Slightly skilled | 37 | 14.3 |  |  |
| Not at all skilled | 8 | 3.1 |  |  |
| No opinion | 7 | 2.7 |  |  |
| Frequency of LLM chatbot use for exercise in past year |  |  |  |  |
| 2-3 times | 70 | 27.1 |  |  |
| 4-5 times | 66 | 25.6 |  |  |
| 6-10 times | 40 | 15.5 |  |  |
| More than 10 times | 82 | 31.8 |  |  |
| LLM chatbot used most for exercise |  |  |  |  |
| ChatGPT | 175 | 67.8 |  |  |
| Gemini | 47 | 18.2 |  |  |
| Co-Pilot | 18 | 7.0 |  |  |
| Other | 18 | 7.0 |  |  |
| Length of use of LLM chatbot for exercise |  |  |  |  |
| Past month | 49 | 19.0 |  |  |
| 1-6 months | 63 | 24.4 |  |  |
| 7-12 months | 70 | 27.1 |  |  |
| 1-2 years | 61 | 23.6 |  |  |
| More than 2 years ago | 15 | 5.8 |  |  |
| Has paid subscription for LLM chatbot | 64 | 24.8 |  |  |
| Has shared weight/BMI with LLM chatbot | 111 | 43.0 |  |  |
| Has shared other lab values/measures with LLM chatbot | 74 | 28.7 |  |  |
| Perceived relevance of LLM chatbot for exercise<sup>a</sup> |  |  | 4.16 | 0.60 |
| Perceived accuracy of LLM chatbot for exercise<sup>a</sup> |  |  | 4.09 | 0.68 |
| Perceived trustworthiness of LLM chatbot for exercise<sup>a</sup> |  |  | 4.03 | 0.69 |
| Perceived usefulness of LLM chatbot for exercise<sup>a</sup> |  |  | 4.16 | 0.60 |
| Emotional bond with LLM chatbot<sup>a</sup> |  |  | 2.96 | 1.29 |
| Report increase in minutes/week of exercise | 140 | 54.3 |  |  |
| Report change in type of exercise | 175 | 67.8 |  |  |

a. scale range: 1-5, with higher values indicating higher level of construct.

### Characteristics of LLM chatbot use for exercise

Participants varied in their extent of LLM chatbot use for exercise, with the majority (52.7%) reporting using it 5 or fewer times in the past year, while nearly a third of participants (31.8%) report 10 or more uses in the past year (see Table 2). ChatGPT was the most popular LLM chatbot, with 67.8% endorsing it as their most used LLM chatbot for exercise topics.

Nearly a quarter of participants (24.8%) reported that they paid for a subscription to an LLM chatbot that they use for exercise topics. Participants had been using LLM chatbots for exercise for varying amounts of time, with 19% reporting their first use for exercise topics only in the past month, while 29.5% reported using it for more than a year.

Participants reported a range of topics and purposes for LLM use, with the most endorsed including making an exercise plan (53.1%), nutrition related to exercise (51.2%), advice on amount of exercise to do (47.3%), specific exercises to try (46.5%) and use for motivation or emotional support (43.4%; See table 3). Approximately 43% of participants reported entering weight or BMI into an LLM chatbot, while 28.7% reported sharing specific lab values or other health measures (beyond weight/BMI).

**Table 3.** Endorsement of specific topics of LLM chatbot for exercise, nutrition, and weight-related topics among recent users of LLM chatbots for exercise (n=258).

|  | N | % |
| --- | --- | --- |
| **Exercise-related topics** |  |  |
| Making an exercise plan/routine | 137 | 53.1 |
| Nutrition related to exercise | 132 | 51.2 |
| Amount of exercise to do | 122 | 47.3 |
| Getting ideas for specific exercises to try | 120 | 46.5 |
| Exercise motivation or emotional support | 112 | 43.4 |
| How to perform specific exercise | 107 | 41.5 |
| Exercising to lose weight | 99 | 38.4 |
| Calculating things related to exercise | 94 | 36.4 |
| Exercise modifications (e.g., for injury) | 83 | 32.2 |
| Explain exercise topics | 73 | 28.3 |
| Help meeting specific goal (e.g., preparing for race) | 53 | 20.5 |
| **Nutrition-related topics** |  |  |
| Asked any nutrition questions | 230 | 89.1 |
| Recipes | 122 | 47.3 |
| Meal planning | 92 | 35.7 |
| Estimate of calories, etc. | 91 | 35.3 |
| Motivation for diet | 91 | 35.3 |
| Nutrition and exercise | 79 | 30.6 |
| For medical conditions | 79 | 30.6 |
| Dietary strategies for weight loss | 74 | 28.7 |
| Recipe moderation | 69 | 26.7 |
| Creating grocery list | 62 | 24.0 |
| Specific dietary approaches | 53 | 20.5 |
| **Weight loss-related topics** |  |  |
| General weight loss strategies | 98 | 38.0 |
| Non-prescription weight loss substances | 68 | 26.4 |
| Bariatric surgery | 20 | 7.8 |
| **Weight loss medication-related topics** |  |  |
| Medications for weight loss (general) | 51 | 19.8 |
| Side effects and risks of medications | 33 | 12.8 |
| How much weight loss to expect | 26 | 10.1 |
| Cost or insurance coverage | 24 | 9.3 |
| How medication works | 23 | 8.9 |
| How to get medications | 19 | 7.4 |
| Comparing medications | 19 | 7.4 |

### Effects of LLM chatbot use

The majority of participants (54.3%; n=140) reported that their amount of exercise had increased as a result of using an LLM chatbot among those reporting an increase, the mean increase was 55.6 minutes per week (SD=55.6). A larger proportion (67.8%; n=175) reported that LLM chatbot use had changed the type of exercise they engaged in. Open-ended responses revealed that the most commonly reported changes related to trying new exercises (n=26); change or new use of equipment for exercise (n=26); increasing or changing strength training in general (n=24); increasing or changing strength training focused on a specific body part (n=24); changing or improving exercise form (n=13); stretching (n=13); and tailoring exercise to individual characteristics, such as pain and injuries (n=11). Less frequent but notable responses included increasing variety (n=5), addressing psychological factors (n=4; e.g., enjoyment of exercise), changing exercise environment (n=4), changing approach to resting in relation to exercise (n=2), and integrating progress tracking (n=1). See Supplemental text for full data.

A minority of participants reported a change in use of exercise-related resources related to their use of LLM chatbots, with some participants reporting an increase and some a decrease, as shown in Table 4. Approximately a quarter of participants reported greater use of gym/workout studio (26.4%), wearable technology (23.3%), and exercise-focused discussions with healthcare providers (25.6%), whereas only about 10% reported a decrease in use of those resources. Other resources, such as a personal trainer/coach and exercise apps were evenly split between participants reporting an increase in use and a decrease in use related to LLM chatbot use.

**Table 4.** Change in use of physical activity-related resources with use of LLM chatbot.

|  | Use more |  | Use less |  | No change |  |
| --- | --- | --- | --- | --- | --- | --- |
|  | N | % | N | % | N | % |
| Gym/ workout studio | 68 | 26.4 | 25 | 9.7 | 165 | 64.0 |
| Discuss exercise with healthcare provider | 66 | 25.6 | 30 | 11.6 | 162 | 62.8 |
| Personal trainer or coach | 40 | 15.5 | 45 | 17.4 | 173 | 67.1 |
| Other fitness facilities | 46 | 17.8 | 31 | 12.0 | 181 | 70.2 |
| YouTube videos for exercise | 96 | 37.2 | 42 | 16.3 | 120 | 46.5 |
| Google search engine | 84 | 32.6 | 84 | 32.6 | 90 | 34.9 |
| Exercise apps | 63 | 24.4 | 54 | 20.9 | 141 | 54.7 |
| Wearable technology | 60 | 23.3 | 27 | 10.5 | 171 | 66.3 |
| Social media/influencer content | 61 | 23.6 | 50 | 19.4 | 147 | 57.0 |

### Differences between those who did and did not increase exercise

In exploratory analyses, participants who reported that they had increased their exercise were compared to those who did not (see Table 5). Those who reported an LLM chatbot-related boost in exercise were on average younger, reported more frequent general LLM chatbot use, had greater perceptions of LLM chatbots’ accuracy and trustworthiness, a higher emotional bond with LLM chatbots, and were more likely to use an LLM chatbot for motivational or emotional support for exercise. We focus here on motivation/emotional support because it was among the largest association with reported exercise increase; several additional exercise-related topics were also significantly associated with reported increase and analysis of all topics as it relates to exercise change are provided in Supplementary Material.

**Table 5.** Differences between those who did and did not report increased physical activity.

| | Increased<br>Min/week | | Did not<br>increase | | $\chi^2/t$ | p-value |
| --- | --- | --- | --- | --- | --- | --- |
|  | N/M | %/SD | N/M | %/SD |  |  |
| Gender |  |  |  |  | 0.12 | 0.73 |
| Male | 73 | 52.9% | 65 | 47.1% |  |  |
| Female | 67 | 55.8% | 53 | 44.2% |  |  |
| Age | 39.8 |  | 43.9 |  | 2.18 | 0.03 |
| LLM use frequency (general) |  |  |  |  | 5.81 | 0.0546 |
| Once a week or less | 22 | 40% | 33 | 60% |  |  |
| Several times a week | 51 | 59.3% | 35 | 40.7% |  |  |
| Daily or more | 67 | 57.3% | 50 | 42.7% |  |  |
| Amount of LLM use for exercise |  |  |  |  | 2.60 | 0.11 |
| <10 x in past year | 89 | 50.6% | 87 | 49.4% |  |  |
| >10x in past year | 51 | 62.2% | 31 | 37.8% |  |  |
| Length of use |  |  |  |  | 5.32 | 0.07 |
| >6 month | 69 | 61.6% | 43 | 38.4% |  |  |
| 7-12 months | 31 | 44.3% | 39 | 55.7% |  |  |
| >12 months | 40 | 52.6% | 36 | 47.4% |  |  |
| Perceived relevance | 4.22 | 0.64 | 4.09 | 0.55 | 1.77 | 0.08 |
| Perceived accuracy | 4.21 | 0.66 | 3.94 | 0.69 | 3.16 | 0.002 |
| Perceived trustworthiness | 4.14 | 0.64 | 3.90 | 0.72 | 2.76 | 0.01 |
| Perceived usefulness | 4.22 | 0.64 | 4.09 | 0.55 | 1.77 | 0.08 |
| Emotional bond | 3.24 | 1.26 | 2.63 | 1.26 | 3.91 | <0.001 |
| Report use LLM for motivation |  |  |  |  | 19.98 | <0.0001 |
| Yes | 79 | 70.5% | 33 | 29.5% |  |  |
| No | 61 | 41.8% | 85 | 58.2% |  |  |

### Engagement with nutrition and weight related content

Use of LLM chatbots for nutrition topics were endorsed by 89.1% of participants, with the most endorsed topics being recipes, estimates of calories or other nutrition, meal planning, and motivation for dietary change (see Table 5). Use of LLM chatbots for general weight loss strategy topics were endorsed by 38.0% of the sample, for non-prescription weight loss substances by 26.4%, for weight loss medications by 19.8%, and for bariatric surgery by 7.8%. Among those querying about weight loss medication, side effects (12.8% of all participants) and amount of weight loss with medication (10.1%) were the most common topics.

## Discussion

Individuals are increasingly turning to LLM chatbots for answers and support related to health topics, including exercise and nutrition.^5,6,23^ In this study, we surveyed US adults who have recently engaged with LLM chatbots for exercise topics in order to understand how they are using these tools and their perceptions of the effects of use. Respondents reported receiving LLM chatbot assistance for a variety of exercise-related topics, most commonly exercise planning, exercise-related nutrition, and motivational or emotional support. The majority of participants reported that LLM chatbot use had increased their weekly exercise and changed the types of exercises they performed. In exploratory analyses, those who reported increases in exercise with LLM chatbot use had greater perceptions of LLM chatbot accuracy and trustworthiness, a stronger emotional bond with the chatbot, and were more likely to use the chatbot for motivational or emotional support for exercise.

This survey is the first, to our knowledge, to characterize how U.S. adults are using LLM chatbots for exercise-related purposes and to document the widespread perception that these tools are influencing both exercise amount and the type of exercise engagement. These findings suggest the potential for commercial, freely available LLM chatbots to be a valuable contributor to the promotion of exercise. However, because these data are cross-sectional and self-reported, they should be interpreted as evidence of perceived impact rather than a casual effect. More rigorous causal designs, such as randomized trials or quasi-experimental designs, are needed to determine whether LLM chatbot use changes exercise behavior and under what conditions.

While more rigorous causal research is needed, prior research speaks to the potential of LLM chatbots to affect health behaviors. General-purpose LLM chatbots have shown promise for changing attitudes towards certain health behaviors, including increasing vaccination intentions^24^ and reducing belief in health-related conspiracy theories.^25^ In a study where participants were suggested to use ChatGPT for two weeks, qualitative interviews show that some found it helpful in improving their health behavior, including physical activity.^26^ Distinct from general-purpose LLMs, research and commercial teams have developed specialized LLMs (i.e., LLMs trained for a specific purpose using specialized source text and/or adjusting model characteristics). These LLM chatbots have begun to be tested and show potential; for example a recently completed trial showed success in reduction of depression^27^ and a pilot trial has shown promise in smoking cessation.^28^ Physical activity focused specialized LLM chatbots have also begun to be developed and evaluated, ^29,30^ though the work is still in early stages.

While specialized LLMs are likely to have an important role in exercise promotion, many individuals will continue to seek out and use the tools that are most accessible and most familiar—including general purpose LLMs that are free and publicly available. If future studies support the beneficial effects of use of general LLM chatbots for increasing exercise, use of these tools offers a new opportunity to reach individuals who might otherwise have limited access to support and information to help them become more active. Given the shortage of qualified healthcare professionals, LLM chatbots may be a valuable supplement for individuals to obtain information and support for exercise. At the same time, there are clear risks for seeking health information from chatbots.^31^ Further research is needed to identify potential harms of LLM chatbot use for physical activity. These findings also motivate development of more structured, safety-aware conversational systems for health behavior support, including domain-adapted systems that can provide grounded, explanation-oriented, and context-sensitive guidance.

Notably, in exploratory analyses, those who reported increased exercise with LLM use differed from those who had not reported increased exercise in their perceptions and use of LLMs. Participants reporting an increase in exercise reported higher levels of trust and perceptions of chatbot accuracy. These participants also reported a higher emotional bond with the tool and were more likely to use the LLM chatbots for motivational or emotional support for exercise. As in other intervention formats,^32,33^ having a sense of connection and trust with the source of support, and addressing motivation and other psychosocial drivers of behavior, are likely important for behavior change with LLM chatbots.

Participants also reported changes in use of other exercise-related resources, but the pattern did not suggest simple displacement. Interestingly, a quarter of respondents reported asking more questions about exercise of their healthcare providers since starting LLM chatbot use for exercise, whereas half as many reported asking fewer questions. This suggests that, for most users, LLM chatbot use for exercise topics will not displace obtaining of guidance from healthcare professionals. Notably, the use of personal trainers was more balanced between those who reported using more (15.5%) and those reporting less use (17.4%). Potentially, the rise of LLM chatbots for exercise topics may change the experience of personal training (e.g., through integration of LLM chatbot information into sessions) but not substantially reduce overall demand for this service.

### Strengths and Limitations

This study has a number of limitations. First, the sample may not be representative of the target population. To date, there is no nationally representative sample indicating what percentage of the US population uses LLMs for exercise. However, the broader screening sample was approximately representative of the U.S. population with respect to gender and racial composition, which provides some reassurance that the analytic sample is not drawn from a highly idiosyncratic subgroup of respondents. An additional important limitation is that, as a cross-sectional survey, the data on behavior change are subject to self-report biases. Another limitation is that we did not have validated measures for all constructs of interest given the novelty of the topic area; however, we evaluated new items through pilot testing prior to deploying the survey. Additionally, our comparison between those who did and did not exercise more with LLM chatbot use was exploratory and not pre-registered, and our sample size may have limited our ability to detect smaller but still meaningful differences between those who did and did not exercise more.

This study also has numerous strengths. Our sample had diversity with regard to age, gender, race/ethnicity, and other demographic factors. Additionally, we used a variety of best practices to avoid selection effects and fraudulent or bot responses and surveyed a national sample. Our survey items were pilot tested by the target population to ensure comprehensibility and comprehensiveness.

## Conclusions

To our knowledge, this is the first study to characterize how individuals are using LLM chatbots for exercise-related purposes and to document perceived effects on exercise amount, exercise type, and use of other exercise-related resources. The findings suggest that LLM chatbots may already be influencing exercise-related behavior for some users, particularly when they are used in ways that users perceive as trustworthy, accurate, and motivationally supportive. Future work should test these relationships using causal designs and should inform development of more structured, domain-adapted systems for safe and effective health behavior support.

## Data Availability

The deidentified data set and analytic code is available at the OSF project website.

https://osf.io/rd9hq/overview?view_only=ad35b7b6c111498db0da83276c1e1fef

## Acknowledgements

This work was funded by the University of Florida Department of Health and Human Performance. We thank Meena Shankar for contributing to the pilot testing of this survey.

## Conflicts of Interest

Dr. Amy Sheer reports speaking roles with Novo Nordisk and Eli Lilly. The remaining authors do not have any conflicts to report.

## Notes

### Competing Interest Statement

Co-Author Dr. Amy Sheer reports speaking roles with Novo Nordisk and Eli Lilly. The remaining authors do not have any conflicts to report.

### Author Declarations

The University of Florida Institutional Review Board (IRB) - 01 approved of this research.

